# Comparing Midline and Peripherally Inserted Central Catheters – a Randomized Feasibility Trial

**DOI:** 10.1101/2024.06.07.24308509

**Authors:** Alina G. Burek, Kelsey Porada, Matthew R Plunk, Sarah Corey Bauer, Melodee Liegl, Amy Pan, Kathryn E Flynn, David C Brousseau, Reiner Gedeit, Amanda J. Ullman

**Affiliations:** Children’s Wisconsin, 8915 W Connell Ct, Milwaukee, WI, United States; Department of Pediatrics, Medical College of Wisconsin, 8701 W Watertown Plank Rd, Milwaukee, WI, United States; Department of Pediatrics, Nemours Children’s Health Delaware and the Sidney Kimmel Medical College at Thomas Jefferson University; School of Nursing, Midwifery and Social Work, The University of Queensland, Brisbane, QLD, Australia; Children’s Health Queensland Hospital and Health Service, Brisbane, QLD, Australia

## Abstract

**Objectives:** The most effective use of midline catheters in children is not well understood. We aimed to test the feasibility of a trial comparing peripherally inserted central catheters (PICCs) to midline catheters in hospitalized children in need of durable vascular access.

**Methods:** Our study combined a single site, randomized controlled feasibility trial and a prospective observational study comparing PICCs to midline catheters. Hospitalized children ages 2-17 years old in need of non-central, medium-term vascular access (5-14 days) were enrolled for one year; enrollment goal of 30 participants/trial arm. The primary outcome was a four-measure feasibility outcome. Secondary outcomes included time-to-device removal and all-cause failure. Multi-method approaches explored patient/family experience.

**Results:** Between 8/2022-8/2023, only 43 of 260 screened patients met eligibility criteria due to a decrease in eligible PICCs used at our site. A total of 35 patients were enrolled: 8/10 in the trial (4 in each arm) and 27/33 in the observational study (21 midline catheters, 6 PICCs). Our trial eligibility goal was not met. The other feasibility measures were met (n=10): (1) 80% of eligible patients enrolled; (2) 100% received the assigned intervention; (3) 96% of catheter inserters found the study acceptable; (4) no missing data.

**Conclusions:** Due to a decrease in PICC use for non-central, medium-term vascular access needs, a trial comparing devices may not be a practical way to assess the effective use of midline catheters in hospitalized children. Next steps may include an implementation-based study evaluating an intravenous catheter selection algorithm that incorporates midline catheters.

## INTRODUCTION

Most hospitalized children require an intravenous (IV) catheter for delivery of IV fluids and antibiotics.^1^ With the broad range of IV catheters available, and their different profiles of complications,^2^ clinicians need evidence to guide the selection of the most appropriate device. Appropriate IV catheter selection improves treatment efficiency and decreases catheter-related complications.^1^ However, there is lack of evidence-based recommendations regarding effectiveness and safety of certain IV catheters when used in children.^1,3^

Peripherally Inserted Central Catheters (PICCs) are frequently used in hospitalized children for medium (5-14 days) and long-term access (>14 days) and to administer solutions not compatible with peripheral infusion; however, concerns regarding potential inappropriate use of PICCs have been described.^1,4–6^ PICCs are associated with high rates of serious complications like central line-associated blood stream infections (CLABSI) and venous thromboembolism (VTE).

One method of preventing PICC complications and patient harm is to prevent inappropriate placement of PICCs when central venous access is not required, and anticipated length of therapy is <14 days.^3,6^ To reduce inappropriate PICC utilization, alternatives for medium-term, non-central venous access like midline catheters (MC) need to be better understood in children. MCs are peripheral vascular access devices, typically 6-15 cm in length, placed in the upper extremity with the tip ending at or below the axilla.^7,8^ MCs have been recently adopted by some hospitals for short and medium-term venous access (<14 days) due to potential for fewer complications compared to PICCs.^5,9,10^ However, there is a scarcity of literature evaluating the effectiveness of MCs in pediatrics and studies comparing PICCs to MCs in the general pediatric population are lacking.^7^

The purpose of this study was to test the feasibility of a trial comparing PICCs to MCs in hospitalized pediatric patients needing non-central, medium-term vascular access and generate preliminary comparative effectiveness data of the two devices.

## METHODS

### Study Design

We conducted a parallel group pragmatic pilot randomized controlled trial (RCT, *arms 1* and *2*) assessing the feasibility of comparing PICCs to MCs for peripherally compatible therapies of medium-term duration (5-14 anticipated days) in children. When randomization was not possible (e.g., after hours, attending physician did not approve), patients eligible for the study were enrolled in PICC or MC prospective observational only arms (*arms 3* and *4*, Figure 1). The observational arms, specifically the alternative pathway, *arm 4*, was designed as an alternative way of capturing effectiveness data if local practice changed toward early adoption of MCs and decrease in PICC use. Both qualitative and quantitative methods were used to describe patient-reported outcomes in the randomized study arms. The trial was prospectively registered at https://clinicaltrials.gov/ (NCT05346406), and the results reported in accordance with the Consolidated Standard of Reporting Trials (CONSORT) Statement.^11,12^ The local institutional review board approved this study prior to commencement (1853701-3).

**Figure 1:**
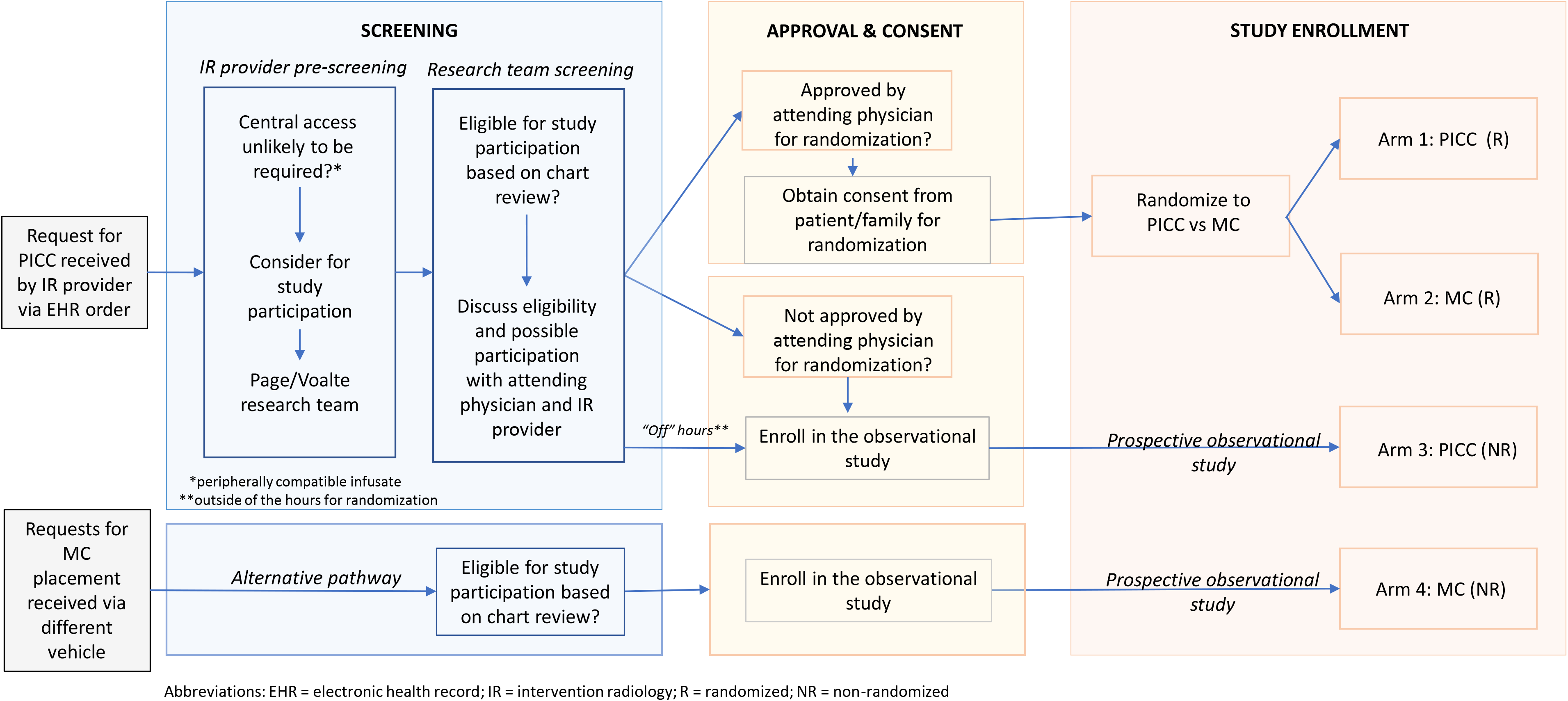
Clinical Trial Protocol – Comparing Peripherally Inserted Central Catheters (PICCs) to Midline Catheters (MCs) – a Feasibility Trial Abbreviations: EHR = electronic health record; IR = intervention radiology; R = randomized; NR = non-randomized

### Setting and Participants

The study took place at a medium-sized pediatric quaternary referral hospital in the Midwest, screening and enrolling Monday through Friday 8-5pm from 8/2022 to 8/2023. <u>Inclusion criteria:</u> patients age 2 to 17 years admitted to the hospital and the medical team requesting placement of a PICC (or MC, alternative pathway) for: (1) anticipated length of IV treatment of 5-14 days, (2) planning only peripherally compatible infusates,^13^ and (3) device not needed at discharge. <u>Exclusion criteria:</u> non-English-speaking family without translators readily available, active bacteremia or VTE at site where device would be placed, urgent need of vascular access (<4 hours), need for more than one lumen, or another central venous catheter already in place.

### Intervention

After recruitment and consent, patients in *arms 1 and 2* were randomized 1:1 to receive:

1. PICC: An uncoated polyurethane PICC, with an external clamp, placed in upper arm, available sizes 3-6Fr (Bard [Becton Dickinson]. Catheter type and size were selected based on interventional radiology (IR) standard protocol.
2. MC: An uncoated MC, available sizes 8cm, 20-22G; Powerglide [Becton Dickinson]. Catheter size was selected by IR or vascular access clinician based on patient size. All MCs were placed in the upper arm with the tip of the catheter ending at or below the axilla.^8^

The patients who could not be randomized (*arms 3 and 4*) received PICC or MC based on the attending physician’s clinical decision.

### Outcomes measures and other variables

#### Feasibility outcomes

The primary outcome applied to the RCT only and was a feasibility outcome determined *a priori* as a composite of four feasibility measures: (1) > 70% of eligible patients agree to enrollment and randomization, (2) > 80% of enrolled patients receive the assigned intervention, (3) > 80% of providers involved in insertion of the device find the study acceptable, and (4) < 5% of data for effectiveness outcomes were missing.^14–16^ Acceptability of the study was defined as a score of 7 or higher given to the question regarding ease/difficulty of device placement (10 the best and 0 the worse placement experience). We aimed to enroll 30 patients in each study group.^14^

#### Effectiveness outcomes

The effectiveness outcomes were time-to-device removal for all reasons (both secondary to completion of therapy and to complications), completion of therapy with initial device, all-cause failure rate, composite complication rate, need for sedation, blood drawing abilities, and additional IV catheters (to complete therapy). Composite complication rate was defined as the rate of any complications, including suspected VTE (ultrasound obtained), suspected CLABSI (blood culture obtained and negative), confirmed VTE (ultrasound positive for clot at catheter site), confirmed CLABSI (defined using the National Healthcare Safety Network surveillance definition^17^), dislodgement, occlusion, phlebitis, line dysfunction, infiltration, leaking. Blood drawing abilities explored were at least one successful blood draw, number of blood draws during length of catheter and lose of blood drawing ability.

Other measures collected included patient demographics (age, gender, race/ethnicity), clinical factors (reason for PICC/MC request, therapy to be infused, requesting service, location, primary diagnosis, complex chronic conditions), IV catheter data (catheter size, insertion site/vessel, number of attempts, inserter type).

#### Patient-reported outcomes

##### Quantitative

Patients and/or guardians enrolled in the RCT were asked to rate procedural pain, anxiety with IV catheter placement, and overall experience. Procedural pain with non-sedated device placement was assessed in patients age >5 years within 24 hours of insertion using the Faces Pain Score (FPS).^18^ Patient/guardian anxiety was assessed pre- and post-device placement using the short State-trait anxiety inventory (STAI) that is validated in children 5 and older.^19^ The score range for the STAI tool is 6 to 24, with 6 points signifying no anxiety and 24 signifying the highest level of anxiety. Overall experience with device placement and maintenance was assessed on a 0-10 scale (0= lowest, 10=highest).^16^

##### Qualitative

We performed semi-structured interviews with the study participants (guardian +/- patient) enrolled in the RCT. Interviews were completed within the week following completion of therapy, in person, prior to discharge. The purpose of the interview was to understand the patient/family’s experience with IV catheter insertion and maintenance during the time of the study.^20,21^ Their advice on ways to improve the process of insertion/maintenance was also collected. (Supplemental Fig.1)

### Study Procedures

Screening of hospitalized children in need of a PICC (or MC for *arm 4*) was completed using our electronic health record (EHR), Epic (Verona, WI). To efficiently identify eligible patients, a Patient List column was created to indicate which patients in the hospital had a PICC order and a separate column to indicate if a catheter was already in place (Supplemental Fig.2). A screening question was added to the PICC order to aid in determining patient eligibility (Supplemental Fig.3). Additionally, IR providers screened the PICC requests and contacted the research team with possible participants. After a candidate was identified, the enrollment criteria were applied to determine eligibility, which was confirmed by the primary attending physician. Informed consent and assent (if patient >7 years old) were obtained prior to enrollment. All IV catheters were placed by trained inserters with ultrasound guidance. Insertion, care and maintenance of all IV catheters followed hospital policy and international guidelines instituted by the hospital.^8^

### Randomization and Blinding

Qualifying RCT participants were randomly allocated 1:1 to *arm 1* or *2*. Stratified block randomization with randomly selected block sizes (2, 4, 6) was used with age category (2-11 years vs 12-17 years) and severity of illness (critical vs acute care) as stratification factors. The randomization sequence was generated using PASS 2023 software (NCSS, LLC. Kaysville, Utah, USA). Due to the clear differences in device appearance and placement modality, masking providers or patients/families to the type of device inserted was not possible.

### Statistical Analysis

#### Sample size calculation

With a goal sample size of 60, we were powered to test feasibility by estimating a participation rate of 70% (eligible patients agree to enrollment and randomization) within a 95% confidence interval of +/- 12%. The study was not powered to detect statistical significance in effectiveness outcomes.

Feasibility outcomes were measured and reported descriptively for the RCT only. Study data were collected using REDCap.^22^ Effectiveness and safety outcomes of RCT arms were summarized separately and in combination with the observational only arms. Outcomes and explored clinical factors were reported as n (%) or median and interquartile range (IQR). SPSS version 28 (IBM Corp., Armonk, NY) was used to analyze the data. Interviews were audio-recorded and transcribed verbatim. Responses were categorized into similar themes.

## RESULTS

### Study participants and device characteristics

Of 260 patients screened over the one-year study period, 43 qualified for enrollment and 35 total patients were enrolled – 8 in the RCT (4 in each arm) and 27 in the prospective observational study (21 MCs and 6 PICCs), see Figure 2. Of the 217 patients with PICC orders who did not qualify for the study, 145 (67%) had 2 or more of the exclusion criteria. Of the 72 (33%) patients screened who had one exclusion criterion, the most common exclusion was need for central access due to infusate properties or anticipated length of therapy >14 days.

**Figure 2:**
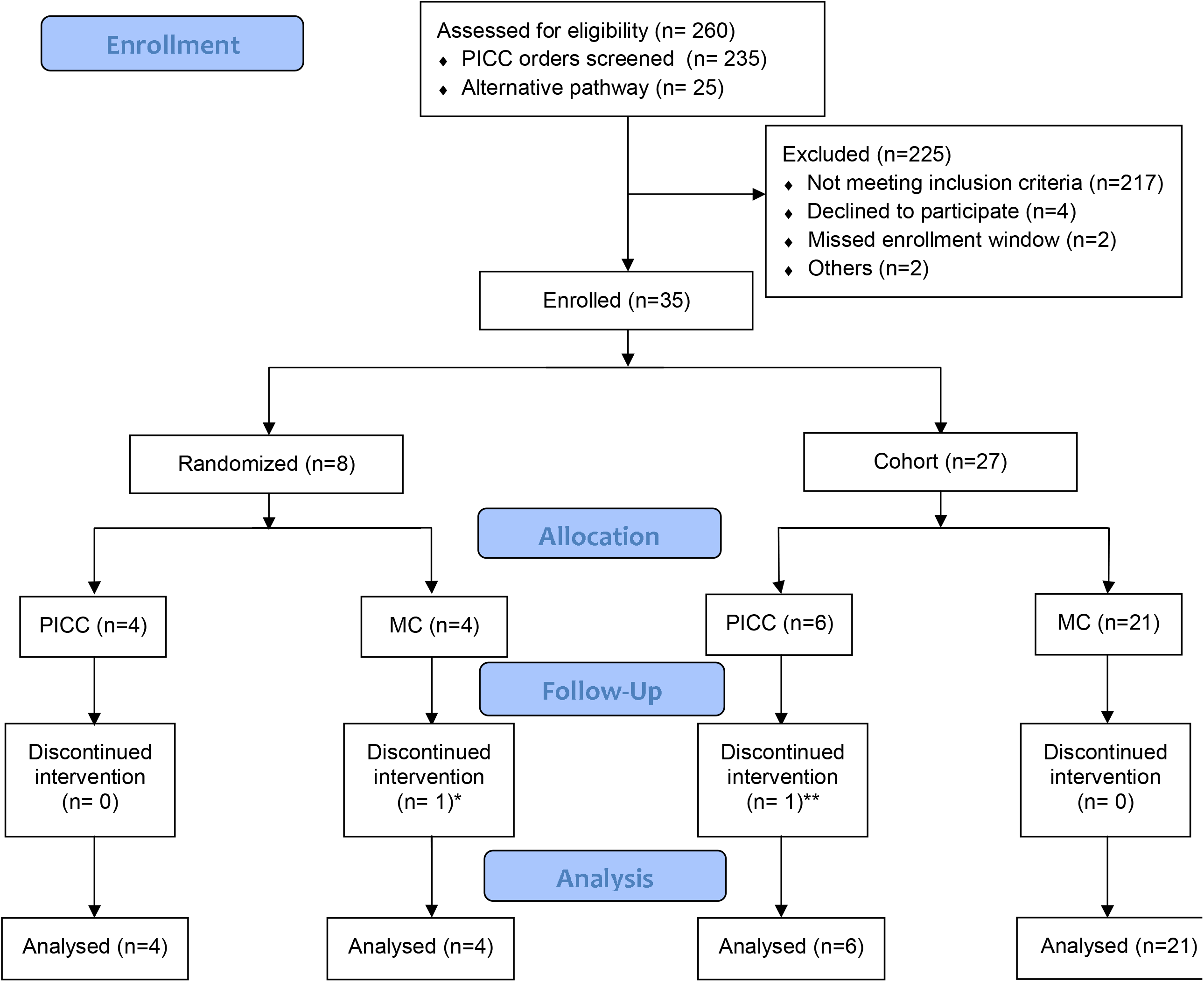
CONSORT Flow Diagram. *Discharged with IV antibiotics and MC rewired to a PICC **Clinically decompensated and required placement of double lumen central venous catheter

Within the cohort, median (IQR) patient age was 9.8 (6.2-15.4), and 57% of patients enrolled were male; 77% had a chronic complex medical condition. Seventy one percent of participants were treated on the acute care units at time of IV catheter request. Neurosurgery was the service for 57% of all IV catheter placements and 90% (18/20) of these were MCs (Table 1).

**Table 1:** Patient and device characteristics by study arm (R – randomized, NR – non-randomized or observational). Data reported as n (%) or median (IQR).

|  | Total (n=35) | Arm 1 (n=4)<br>PICC (R) | Arm 2 (n=4)<br>MC (R) | Arm 3 (n=6)<br>PICC (NR) | Arm 4 (n=21)<br>MC (NR) |
| --- | --- | --- | --- | --- | --- |
| **Patient characteristics** |  |  |  |  |  |
| Age, years | 9.8 (6.2-15.4) | 10.2 (4.0-16.8) | 10.8 (4.5-16.3) | 8.4 (6.2-15.8) | 11.2 (7.7-15.0) |
| Male sex | 20 (57) | 0 ( 0) | 4 (100) | 4 (67) | 12 (57) |
| Race<sup>a</sup> |  |  |  |  |  |
| American Indian /Alaska Native | 1 ( 3) | 0 ( 0) | 0 ( 0) | 0 ( 0) | 1 ( 5) |
| African American | 3 ( 9) | 0 ( 0) | 1 (25) | 0 ( 0) | 2 (10) |
| Caucasian | 29 (88) | 3 (100) | 3 (75) | 6 (100) | 17 (85) |
| Ethnicity<sup>b</sup> |  |  |  |  |  |
| Hispanic | 4 (12) | 0 ( 0) | 2 (50) | 0 ( 0) | 2 (10) |
| Non-Hispanic | 30 (88) | 3 (100) | 2 (50) | 6 (100) | 19 (90) |
| Severity/Location |  |  |  |  |  |
| Acute Care | 25 (71) | 4 (100) | 4 (100) | 6 (100) | 11 (52) |
| Critical Care | 10 (29) | 0 ( 0) | 0 ( 0) | 0 ( 0) | 10 (48) |
| Primary Diagnosis (1) |  |  |  |  |  |
| Medical | 17 (49) | 4 (100) | 4 (100) | 4 (67) | 5 (24) |
| Surgical | 18 (51) | 0 ( 0) | 0 ( 0) | 2 (33) | 16 (76) |
| Primary Diagnosis (2) |  |  |  |  |  |
| Infection | 16 (46) | 4 (100) | 3 (75) | 4 (67) | 5 (24) |
| Non-infection | 19 (54) | 0 ( 0) | 1 (25) | 2 (33) | 16 (76) |
| Primary Diagnosis Type |  |  |  |  |  |
| Respiratory failure | 1 ( 3) | 0 ( 0) | 0 ( 0) | 0 ( 0) | 1 ( 5) |
| Cardiopulmonary arrest | 1 ( 3) | 0 ( 0) | 0 ( 0) | 1 (17) | 0 ( 0) |
| Complicated Pneumonia | 8 (22) | 3 (75) | 0 ( 0) | 3 (50) | 2 (10) |
| Orbital Cellulitis | 1 ( 3) | 0 ( 0) | 0 ( 0) | 0 ( 0) | 1 ( 5) |
| Pulmonary exacerbation of CF | 2 ( 6) | 0 ( 0) | 2 (50) | 0 ( 0) | 0 ( 0) |
| Refractory epilepsy surgery | 19 (54) | 0 ( 0) | 1 (25) | 2 (33) | 16 (75) |
| Septic Shock | 2 ( 6) | 1 (25) | 0 ( 0) | 0 ( 0) | 1 ( 5) |
| VP Shunt Infection | 1 ( 3) | 0 ( 0) | 1 (25) | 0 ( 0) | 0 ( 0) |
| Chronic Complex Medical Condition | 27 (77) | 1 (25) | 4 (100) | 4 (67) | 18 (86) |
| **Catheter characteristics** |  |  |  |  |  |
| Reason for Device Placement |  |  |  |  |  |
| Prolonged Antibiotics Course | 14 (40) | 4 (100) | 3 (75) | 4 (67) | 3 (14) |
| Durable Access | 21 (60) | 0 ( 0) | 1 (25) | 2 (33) | 18 (86) |
| Therapy to be infused |  |  |  |  |  |
| Antibiotics | 15 (43) | 4 (100) | 3 (75) | 4 (67) | 4 (19) |
| Post-operative care/seizure rescue | 19 (54) | 0 ( 0) | 1 (25) | 2 (33) | 16 (76) |
| Sedation while intubated | 1 ( 3) | 0 ( 0) | 0 ( 0) | 0 ( 0) | 1 ( 5) |
| Requesting service |  |  |  |  |  |
| Hospital Medicine | 11 (31) | 4 (100) | 0 ( 0) | 4 (67) | 3 (14) |
| Pulmonary | 2 ( 6) | 0 ( 0) | 2 (50) | 0 ( 0) | 0 ( 0) |
| Neurosurgery | 20 (57) | 0 ( 0) | 2 (50) | 2 (33) | 16 (76) |
| Critical Care | 2 ( 6) | 0 ( 0) | 0 ( 0) | 0 ( 0) | 2 (10) |
| Insertor Type |  |  |  |  |  |
| IR Attending | 3 ( 8) | 0 ( 0) | 0 ( 0) | 1 (17) | 2 ( 9) |
| IR PA | 24 (69) | 3 (75) | 3 (75) | 4 (66) | 14 (67) |
| ICU NP | 1 ( 3) | 0 ( 0) | 0 ( 0) | 0 ( 0) | 1 ( 5) |
| RN | 4 (11) | 0 ( 0) | 1 (25) | 0 ( 0) | 3 (14) |
| Radiology Resident | 2 ( 6) | 1 (25) | 0 ( 0) | 1 (17) | 0 ( 0) |
| ICU Fellow | 1 ( 3) | 0 ( 0) | 0 ( 0) | 0 ( 0) | 1 ( 5) |
| Insertion site/Vessel |  |  |  |  |  |
| Basilic | 17 (49) | 1 (25) | 2 (50) | 2 (33) | 12 (57) |
| Brachial | 7 (20) | 2 (50) | 0 ( 0) | 3 (50) | 2 (10) |
| Cephalic | 11 (31) | 1 (25) | 2 (50) | 1 (17) | 7 (33) |
| Number of Attempts |  |  |  |  |  |
| 1 | 33 (94) | 4 (100) | 3 (75) | 6 (100) | 20 (95) |
| 2 | 2 ( 6) | 0 ( 0) | 1 (25) | 0 ( 0) | 1 ( 5) |

### Primary outcome: feasibility of RCT

Ten patients were eligible for the RCT over the one-year period, significantly lower than the goal of 60 participants. Within this sample, the four feasibility measures were met: (1) 80% (8/10) of eligible patients agreed to enroll and were randomized; (2) 100% received the assigned intervention; (3) 96% of inserters of the device found the study acceptable, and (4) there were no missing data. The reasons for the two refusals to participate in the RCT were “discomfort with research” and “too much going on already”.

### Effectiveness outcomes

Median time-to-device removal for all reasons was 9.5 (IQR 6.7-13.7) days in the combined PICCs (n=10) versus 7.4 (4.7-8.9) days in the combined MCs (n=25) (Table 2 & Figure 3). All-cause failure rate was 0% for PICCs and 36% for MCs (48.6/1000 catheter days). The most common complications that resulted in MC removal were occlusion, infiltration, and dislodgement; each at a rate of 8% (n=2). Composite complication rate (accounting for complications that did not result in catheter removal and for resource utilization) was similar, in 5/10 (50%) of PICCs and 12/25 (48%) of MCs. Regarding blood drawing abilities, 12/24 (50%) MCs were used for blood draws compared to 9/10 (90%) of PICCs; 11/12 (92%) MCs had at least one successful blood draw and 1/11 MC later lost blood drawing abilities.

**Table 2.** Effectiveness and Safety of Peripherally Inserted Central Catheters (PICC) and Midline Catheters (MC). Data reported as n (%) or median (IQR) unless otherwise specified.

|  | PICC |  |  | MC |  |  |
| --- | --- | --- | --- | --- | --- | --- |
|  | Total PICC<br>(n=10) | R<br>(n=4) | NR<br>(n=6) | Total MC<br>(n=25) | R<br>(n=4) | NR<br>(n=21) |
| Time-to-Device Removal for All Reasons, days | 9.5 (6.7-13.7) | 11.0 (7.8-12.0) | 8.5 (5.5-18.9) | 7.4 (4.7-8.9) | 4.7 (3.3-5.9) | 8.0 (6.2-9.1) |
| Completion of Therapy with Initial Device | 10 (100) | 4 (100) | 6 (100) | 16 (64) | 1 (25) | 15 (71) |
| All-Cause Failure Rate | 0 (0) | 0 (0) | 0 (0) | 9 (36) | 3 (75) | 6 (29) |
| Composite Complication Rate: | 5 (50) | 2 (50) | 3 (50) | 12 (48) | 3 (75) | 8 (38) |
| Complications Resulting in Catheter Early Removal |  |  |  |  |  |  |
| Occlusion | 0 (0) | 0 (0) | 0 (0) | 2 (8) | 1 (20) | 1 (5) |
| Infiltration | 0 (0) | 0 (0) | 0 (0) | 2 (8) | 1 (20) | 1 (5) |
| Dislodgement | 0 (0) | 0 (0) | 0 (0) | 2 (8) | 0 (0) | 2 (10) |
| Pain | 0 (0) | 0 (0) | 0 (0) | 1 (4) | 0 (0) | 1 (5) |
| Phlebitis | 0 (0) | 0 (0) | 0 (0) | 0 (0) | 0 (0) | 0 (0) |
| VTE | 0 (0) | 0 (0) | 0 (0) | 0 (0) | 0 (0) | 0 (0) |
| CLABSI | 0 (0) | 0 (0) | 0 (0) | 0 (0) | 0 (0) | 0 (0) |
| Complications Not Resulting in Catheter Removal* |  |  |  |  |  |  |
| Concern for CLABSI | 4 (40) | 1 (25) | 3 (50) | 5 (20) | 2 (50) | 3 (14) |
| Concern for VTE | 0 (0) | 0 (0) | 0 (0) | 0 (0) | 0 (0) | 0 (0) |
| Other Complications | 3 (30) | 1 (20) | 2 (33) | 3 (12) | 2 (50) | 1 (5) |
| Intermittent/positional Occlusion | 0 (0) | 0 (0) | 0 (0) | 3 (12) | 2 (50) | 1 (5) |
| Occlusion (use of tPA) | 2 (20) | 1 (20) | 1 (17) | 0 (0) | 0 (0) | 0 (0) |
| Catheter Kinking | 1 (10) | 0 (0) | 1 (17) | 0 (0) | 0 (0) | 0 (0) |
| Use of Sedation | 9 (90) | 4 (100) | 5 (83) | 21 (84) | 3 (75) | 18 (86) |
| Line Used for Blood Draws | 9 (90) | 4 (100) | 5 (83) | 12 (50) | 2 (50) | 10 (50) |
| Blood drawing abilities |  |  |  |  |  |  |
| Successful Blood Draw (at least one) | 9 (100) | 4 (100) | 5 (100) | 11 (92) | 2 (100) | 9 (90) |
| Number of Successful Blood Draws | 7.0 (4.0-10.5) | 8.0 (5.8-14.0) | 5.0 (2.0-10.0) | 2.0 (1.0-2.0) | 2 (1,3) | 2.0 (1.0-2.0) |
| Catheter Lost Blood Drawing Abilities | 0 (0) | 0 (0) | 0 (0) | 1 (9) | 1 (50) | 0 (0) |
| Blood Draws from Vein | 0.0 (0-1.5) | 0.0 (0.0-2.3) | 0.0 (0.0-1.3) | 0.5 (0-1.8) | 1 (0,2) | 0.0 (0.0-1.0) |
| Reason for Removal |  |  |  |  |  |  |
| No Longer Medically Indicated | 10 (100) | 4 (100) | 6 (100) | 16 (64) | 1 (25) | 15 (71) |
| Complications | 0 (0) | 0 (0) | 0 (0) | 7 (28) | 2 (50) | 5 (24) |
| Home IV Antibiotics (MC rewired to PICC) | 0 (0) | 0 (0) | 0 (0) | 1 (4) | 1 (25) | 0 (0) |
| Irritability | 0 (0) | 0 (0) | 0 (0) | 1 (4) | 0 (0) | 1 (5) |
| Participants that Required Placement of Additional device(s) | 1 (10) | 0 (0) | 1 (17) | 7 (28) | 3 (75) | 4 (19) |
| Type of Additional Device Placed |  |  |  |  |  |  |
| PICC | 0 (0) | 0 (0) | 0 (0) | 1 (14) | 1 (33) | 0 (0) |
| PIVC | 0 (0) | 0 (0) | 0 (0) | 6 (86) | 2 (67) | 4 (100) |
| Untunneled CVC | 1 (100) | 0 (0) | 1 (100) | 0 (0) | 0 (0) | 0 (0) |
| Mean Anxiety Scores (RCT only) |  |  |  |  |  |  |
| Participant Pre-procedure |  | 13.0 |  |  | 9.5 |  |
| Participant Post-procedure |  | 15.0 |  |  | 13.0 |  |
| Guardian Pre-procedure |  | 14.0 |  |  | 12.0 |  |
| Guardian Post-procedure |  | 13.5 |  |  | 13.8 |  |
| Mean Overall Experience with IV Catheter (RCT only) |  | 7.3 |  |  | 9 |  |

\*Same catheter can have more than one complication; Abbreviations: CLABSI = central line associated blood stream infection, CVC = central venous catheter, PIVC = peripheral IV catheter, tPA = tissue plasminogen activator, VTE = venous thromboembolism, R = randomized, NR = non-randomized or observational

**Figure 3:**
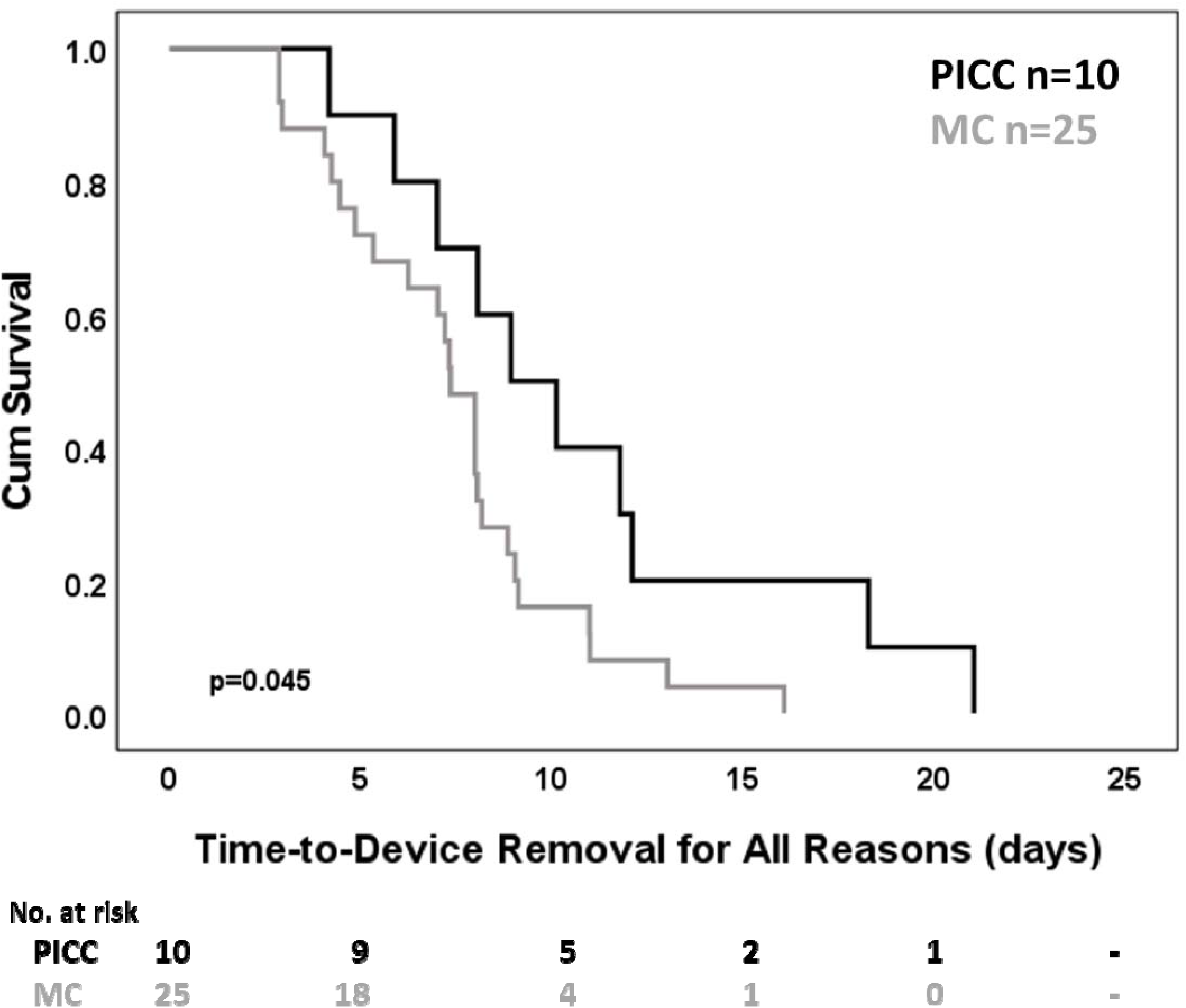
Kaplan Meier survival curve of time-to-device removal by device group

### Patient-reported outcomes (RCT only)

#### Quantitative

Two children out of eight were eligible to provide procedural pain scores, both enrolled in the MC arm; the remaining patients were <5 years old or sedated for the IV catheter placement. Anxiety scores pre- and post-procedure were measured in 4 children and 8 guardians; 2 children and 4 guardians in each arm (Table 2). The mean overall experience score for MCs was 7.3/10 (n=3) and for PICCs was 9/10 (n=4).

#### Qualitative

Seven out of eight guardians or patient-guardian dyads were available for interviews (3 MC, 4 PICC). One guardian could not be reached after multiple attempts. In general, common themes for a positive experience with insertion/maintenance of the IV catheters were sedation for placement (the opposite, no sedation, being a common negative comment); the ability to use the IV catheter for blood draws; the location of the catheter in the upper arm which allowed for better arm movement, better sleep, and decreased visibility. The older participants reported feeling nervous prior to IV catheter placement and prior to its removal – “I was scared because it was like, I’ve never had things in my body before…like a tube inside me” (Participant 9, PICC). Negative comments within the MC arm were related to failed first attempt requiring placement under sedation the following day (toddler participant), occasional discomfort with administration of fluids/medications, complications (e.g., intermittent occlusion). Negative comments in the PICC arm were related to occasional discomfort with administration of fluids/medications and discomfort with dressing changes.

Things that could be improved in the future were use of sedation in younger children (“it took a lot to – for like these nurses to hold him down. So, I don’t know if that happens with other kids, but that’s a lot”; guardian 14, MC); placing an IV catheter that stays in until discharge avoiding multiple peripheral IV catheters (PIVCs); and preparing patients/families about possible complications and the potential need for an additional IV catheter. One guardian reported: “I can recall I think maybe she had three or four IVs before the PICC was finally placed in. Those were the worst for her. She hates having IVs placed. If your study results in fewer net IVs being placed, that’s probably going to help the stress in a lot of children […] (guardian 13, PICC).”

## DISCUSSION

This study aimed to test the feasibility of a trial comparing PICCs to MCs in hospitalized pediatric patients needing non-central, medium-term vascular access, and collect preliminary effectiveness and safety data of the two devices. Our enrollment goal of 30 participants per RCT arm over a one-year period was not met. Even though the other 4 feasibility measures were met, we did not have the goal sample size and, therefore, the statistical power to determine the feasibility of a full-scale effectiveness trial. In fact, the low study eligibility suggests that an effectiveness study is not feasible or practical as currently designed. In a prior retrospective study conducted at our institution, we demonstrated that 139 PICCs placed between 7/2018 and 6/2019 did not meet the requirements necessitating central access (e.g., received peripherally compatible infusate, dwell-time <14 days).^5^ This was the target population for our study. However, PICC use has trended down since 2019,^6^ especially for children requiring medium-term, non-central vascular access, for a variety of reasons. One reason being new recommendations for length of IV antibiotics in children with complicated infections (e.g., bone and joint infections).^23,24^ The new therapeutic option for treatment of pediatric cystic fibrosis, Elexacaftor-Tezacaftor-Ivacaftor, has had a significant impact on the rate of hospitalization for pulmonary exacerbations, with fewer children needing prolonged antibiotics.^25^ Subspecialties like neurosurgery became early adopters of MCs for peri-operative use which is evident in the large representation of neurosurgery patients within our MC observational group. While a powered RCT would be preferable for comparing the effectiveness of PICCs to MCs, due to the decrease in PICC use for non-central, medium-term vascular access needs, a comparative trial randomizing the two devices at the individual level is not practical. Instead, we need to better understand the barriers and facilitators surrounding device selection and clinical decision making across our nation and use a multisite implementation-based design to explore important clinical outcomes (e.g., CLABSI, extravasations). Such a trial could be a hybrid multisite clustered RCT examining the implementation and clinical impact of imbedding an IV catheter selection algorithm that incorporates PICCs and MCs along with other commonly used IV catheters. Due to ethical considerations, RCT entrance required a request for PICC; however, it is likely that many hospitalized children in need of durable access and satisfying all the other inclusion criteria received PIVCs. This is the population with length of stay of 5-14 days that previously may have had a PICC inserted. During our interviews, patients/families report that they would prefer to have one IV catheter from admission to discharge. Based on our preliminary data, MCs may be inferior to PICCs in some ways; however, there is evidence that MCs are superior to PIVCs.^26,27^ Next steps should be taken to identify situations where an MC could be the only IV catheter used to provide therapy, replacing multiple PIVCs per hospitalization.

During the one-year study period, 25/35 children enrolled received an MC. Since placing a PICC would have been the primary option in these children prior to our study, we avoided 24 PICC placements (one participant transitioned from MC to PICC). However, we were not powered to find differences in effectiveness and safety between devices. Because of the sample size and limitations, caution should be taken in data interpretation and its use for clinical decision making. Based on our prospectively collected data from 25 MCs used, we can make general observations. (1) The majority of MCs lasted for ∼7 days, so the population that could benefit from an MC may be children in need of vascular access for 4-9 days. However, 37% of all MCs failed prior to completion of therapy and almost half of those required at least one additional IV catheter, most commonly a PIVC. This failure rate, even though higher than that of PICCs, is superior to that of the PIVC which can be up to 50-60%, with PIVCs lasting for an average of 3.5 days.^26,28^ (2) We have shown that MCs can be used for blood draws; with occlusion being one of the commonly seen complications, ability to draw blood may be lost. The median number of blood draws per MC was 2, so larger studies are needed to determine if MCs could be used in children known to need frequent blood draws (>1/day). (3) The location in the upper arm was highly valued by patients/families, as it allowed for improved arm mobility, better sleep and less accessible to the younger patients. Similarly, patients and families valued placement under sedation, a topic that should be explored further since sedation has additional risks.

Our patient-reported outcome data are consistent with previous reports, that IV catheter placement generates anxiety in children and their families.^29^ The STAI anxiety scores for both patients and guardians were above 6, the standard threshold for a positive anxiety score.

Many challenges may be encountered during enrollment in studies involving hospitalized children. Challenges to consider when designing inpatient RCTs are lack of guardian availability at bedside for consent; last minute clinical decisions without sufficient time left for consent (e.g., the request to place a PICC as an ‘add on’ to a scheduled procedure); the adequacy of approaching families of critically ill children; and unexpected changes in treatment plan (e.g., MC rewired and replaced with a PICC for outpatient IV antibiotics).

Our study has limitations. The low rate of enrollment did not allow us to determine if the feasibility measures would have been accomplished, information that would be valuable for future studies conducted in the inpatient acute care settings. Children <2 years old were excluded due to size incompatibility with the available MC. Only 50% of MCs were used for blood draws in our cohort; we speculate this was primarily due to existing hospital policies that, at the time of the study, did not permit blood draws from peripheral catheters. Even though we had special permission for blood drawing for study participants, communicating this change to all the clinicians involved was challenging. To improve this in the future, EHR lab orders should align with proper collection (e.g., “unit collect” vs “lab collect”). At our institution, MCs are commonly placed awake without sedation with the use of topic anesthetic only; however, as noted in our outcome data, 84% of MCs were placed under sedation due to a large sample of peri-operative cases that received MCs in the operating room. During the feasibility trial, some of the measures included in the initial protocol and determined *a priori* were removed. They include prolonged NPO time > 8 hours (many children were NPO for a different procedure), time-to-placement of IV catheter (sometimes the order was placed after IV catheter insertion), and time-to-last blood draw (too challenging to accurately measure).

Strengths of our study include the innovative study design combining an RCT and observational study, the creative EHR-based screening strategies^30^ and assessment of patient-reported outcomes.

## CONCLUSION

An effectiveness trial comparing PICCs to MCs in hospitalized children needing non-central, medium-term vascular access may not be feasible or practical due to an overall downward trend in PICC use in this population locally and likely nationally. Next steps may include an implementation focused study, which may include cluster-based trial methods to test the implementation contexts and clinical impact of an IV catheter selection algorithm.

## Data Availability

All data produced in the present work are contained in the manuscript

## Notes

### Competing Interest Statement

Dr Ullman's employer (UQ) has received investigator-initiated research grants from vascular access product manufacturers (3M, Becton Dickinson, Medline) on behalf of her research, unrelated to the submitted project. David Brousseau receives consultancy funding from CSL Behring unrelated to this work. The remaining authors have no conflicts of interest to disclose.

### Clinical Trial

ClinicalTrial.gov ID: NCT05346406

### Funding Statement

Dr. Burek received one-year pilot funding for this study from the Children Research Institute, Children Wisconsin, Milwaukee. In addition, her work on this study was in part completed while participating in a 2-year Clinical Research Scholar Program funded by a training grant (UL1 TR001436).

### Author Declarations

The Medical College of Wisconsin's institutional review board approved this study prior to commencement (1853701-3).

## References

1. Paterson RS, Chopra V, Brown E, et al. Selection and insertion of vascular access devices in pediatrics: A systematic review. Pediatrics. 2020;145(June):S243–S268. doi:10.1542/peds.2019-3474H

2. Ullman AJ, Marsh N, Mihala G, Cooke M, Rickard CM. Complications of central venous access devices: A systematic review. Pediatrics. 2015;136(5):e1331–e1344. doi:10.1542/peds.2015-1507

3. Ullman AJ, Bernstein SJ, Brown E, et al. The Michigan appropriateness guide for intravenous catheters in pediatrics: miniMAGIC. Pediatrics. 2020;145(June):S269–S284. doi:10.1542/peds.2019-3474I

4. Gibson C, Connolly BL, Moineddin R, Mahant S, Filipescu D, Amaral JG. Peripherally inserted central catheters: Use at a tertiary care pediatric center. J Vasc Interv Radiol. 2013;24(9). doi:10.1016/j.jvir.2013.04.010

5. Burek AG, Parker J, Bentzien R, Talbert L, Havas M, Hanson SJ. The Development of a Long Peripheral Catheter Program at a Large Pediatric Academic Center: A Pilot Study. Hosp Pediatr. 2020;10(10):897–901. doi:10.1542/hpeds.2020-0181

6. Burek AG, Davis MB, Pechous B, et al. Inappropriate Use of Peripherally Inserted Central Catheters in Pediatrics: A Multisite Study. Hosp Pediatr. 2024;14(3):180–188. doi:10.1542/hpeds.2023-007518

7. Burek AG, Liljestrom T, Dundon M, Shaughnessy EE, Suelzer E, Ullman A. Long peripheral catheters in children: A scoping review. J Hosp Med. 2022;(June):1–10. doi:10.1002/jhm.12968

8. Gorski LA, Hadaway L, Hagle ME, et al. Infusion Therapy Standards of Practice, 8th Edition. J Infus Nurs. 2021;44. doi:10.1097/NAN.0000000000000396

9. Moureau N, Sigl G, Hill M. How to Establish an Effective Midline Program: A Case Study of 2 Hospitals. JAVA - J Assoc Vasc Access. 2015;20(3):179–188. doi:10.1016/j.java.2015.05.001

10. DeVries M, Lee J, Hoffman L. Infection free midline catheter implementation at a community hospital (2 years). Am J Infect Control. 2019;47(9). doi:10.1016/j.ajic.2019.03.001

11. Schulz KF, Altman DC, Moher D. CONSORT 2010 Statement: Updated guidelines for reporting parallel group randomised trials. Ital J Public Health. 2010;7(3). doi:10.4178/epih/e2014029

12. Eldridge SM, Chan CL, Campbell MJ, et al. CONSORT 2010 statement: Extension to randomised pilot and feasibility trials. Pilot Feasibility Stud. 2016;2(1). doi:10.1186/s40814-016-0105-8

13. Clark E, Giambra BK, Hingl J, Doellman D, Tofani B, Johnson N. Reducing risk of harm from extravasation: A 3-tiered evidence-based list of pediatric peripheral intravenous infusates. J Infus Nurs. 2013;36(1). doi:10.1097/NAN.0b013e3182798844

14. Thabane L, Ma J, Chu R, et al. A tutorial on pilot studies: The what, why and how. BMC Med Res Methodol. 2010;10. doi:10.1186/1471-2288-10-1

15. Arnold DM, Burns KEA, Adhikari NKJ, Kho ME, Meade MO, Cook DJ. The design and interpretation of pilot trials in clinical research in critical care. Crit Care Med. 2009;37(SUPPL. 1). doi:10.1097/CCM.0b013e3181920e33

16. Kleidon TM, Schults JA, Wainwright C, et al. Comparison of midline catheters and peripherally inserted central catheters to reduce the need for general anesthesia in children with respiratory disease: A feasibility randomized controlled trial. Pediatr Anesth. 2021;31:985–995. doi:10.1111/pan.14229

17. Centers for Disease Control and Prevention. National Healthcare Safety Network (NHSN) Patient Safety Component Manual.; 2022. https://www.cdc.gov/nhsn

18. Hunter M, McDowell L, Hennessy R, Cassey J. An evaluation of the faces pain scale with young children. J Pain Symptom Manage. 2000;20(2). doi:10.1016/S0885-3924(00)00171-8

19. Nilsson S, Buchholz M, Thunberg G. Assessing Children’s Anxiety Using the Modified Short State-Trait Anxiety Inventory and Talking Mats: A Pilot Study. Nurs Res Pract. 2012;2012. doi:10.1155/2012/932570

20. Larsen E, Keogh S, Marsh N, Rickard C. Experiences of peripheral IV insertion in hospital: A qualitative study. Br J Nurs. 2017;26(19). doi:10.12968/bjon.2017.26.19.S18

21. Sharp R, Grech C, Fielder A, Mikocka-Walus A, Cummings M, Esterman A. The patient experience of a peripherally inserted central catheter (PICC): A qualitative descriptive study. Contemp Nurse. 2014;48(1). doi:10.1080/10376178.2014.11081923

22. Harris PA, Taylor R, Minor BL, et al. The REDCap consortium: Building an international community of software platform partners. J Biomed Inform. 2019;95. doi:10.1016/j.jbi.2019.103208

23. Woods CR, Bradley JS, Chatterjee A, et al. Clinical Practice Guideline by the Pediatric Infectious Diseases Society and the Infectious Diseases Society of America: 2021 Guideline on Diagnosis and Management of Acute Hematogenous Osteomyelitis in Pediatrics. J Pediatric Infect Dis Soc. 2021;10(8). doi:10.1093/jpids/piab027

24. House SA, Hall M, Ralston SL, et al. Development and Use of a Calculator to Measure Pediatric Low-Value Care Delivered in US Children’s Hospitals. JAMA Netw Open. 2021;4(12):1–13. doi:10.1001/jamanetworkopen.2021.35184

25. McNally P, Lester K, Stone G, et al. Improvement in Lung Clearance Index and Chest Computed Tomography Scores with Elexacaftor/Tezacaftor/Ivacaftor Treatment in People with Cystic Fibrosis Aged 12 Years and Older – The RECOVER Trial. Am J Respir Crit Care Med. 2023;208(9). doi:10.1164/rccm.202308-1317OC

26. Qin KR, Ensor N, Barnes R, Englin A, Nataraja RM, Pacilli M. Standard versus long peripheral catheters for multiday IV therapy: A randomized controlled trial. Pediatrics. 2021;147(2):e2020000877. doi:10.1542/peds.2020-000877

27. Kleidon TM, Gibson V, Cattanach P et al. Midline Compared With Peripheral Intravenous Catheters for Therapy of 4 Days or Longer in Pediatric Patients: A Randomized Clinical Trial. JAMA Pediatr.

28. Helm RE, Klausner JD, Klemperer JD, Flint LM, Huang E. Accepted but Unacceptable: Peripheral IV Catheter Failure. J Infus Nurs. 2015;38(3):189–203. doi:10.1097/nan.0000000000000326

29. Sharp R, Muncaster M, Baring CL, Manos J, Kleidon TM, Ullman AJ. The parent, child and young person experience of difficult venous access and recommendations for clinical practice: A qualitative descriptive study. J Clin Nurs. 2023;(October 2022):1–16. doi:10.1111/jocn.16759

30. Chishtie J, Sapiro N, Wiebe N, et al. Use of Epic Electronic Health Record System for Health Care Research: Scoping Review. J Med Internet Res. 2023;25(1). doi:10.2196/51003

