## Supplemental Figure 1 for "Comparing Midline and Peripherally Inserted Central Catheters – a Randomized Feasibility Trial"

**Semi structured interview**

***(At time of line removal/discharge)***

Question 1.

*What was your/your child’s experience of having this PICC/MC inserted?*

*1a. What made it a positive experience?*

*1b. What made it a negative experience?*

Question 2.

*What was your/your child’s experience of having this PICC/MC used for treatment?*

*2a. What made it a positive experience?*

*2b. What made it a negative experience?*

Question 3.

*When you think about your own experience and your child’s experience with the vascular line (placement, maintenance, removal), how would you rate your overall experience?*

*0=worst experience possible and 10=best experience possible*

Question 4.

*Did you child have a PICC/MC placed in the past? If yes:*

*3a. How was this experience different from your previous experiences?*

*3b. Do you prefer a PICC or MC? (if applicable)*

Question 5.

*4a. What makes for a positive vascular device experience?*

*4b. How can insertion of your device be improved?*

*4c. How can ongoing maintenance of your device be improved?*
