## Supplemental Figure 2 for "Comparing Midline and Peripherally Inserted Central Catheters – a Randomized Feasibility Trial"

Supplemental Figure 2: Special Columns for Eligible Patient Identification

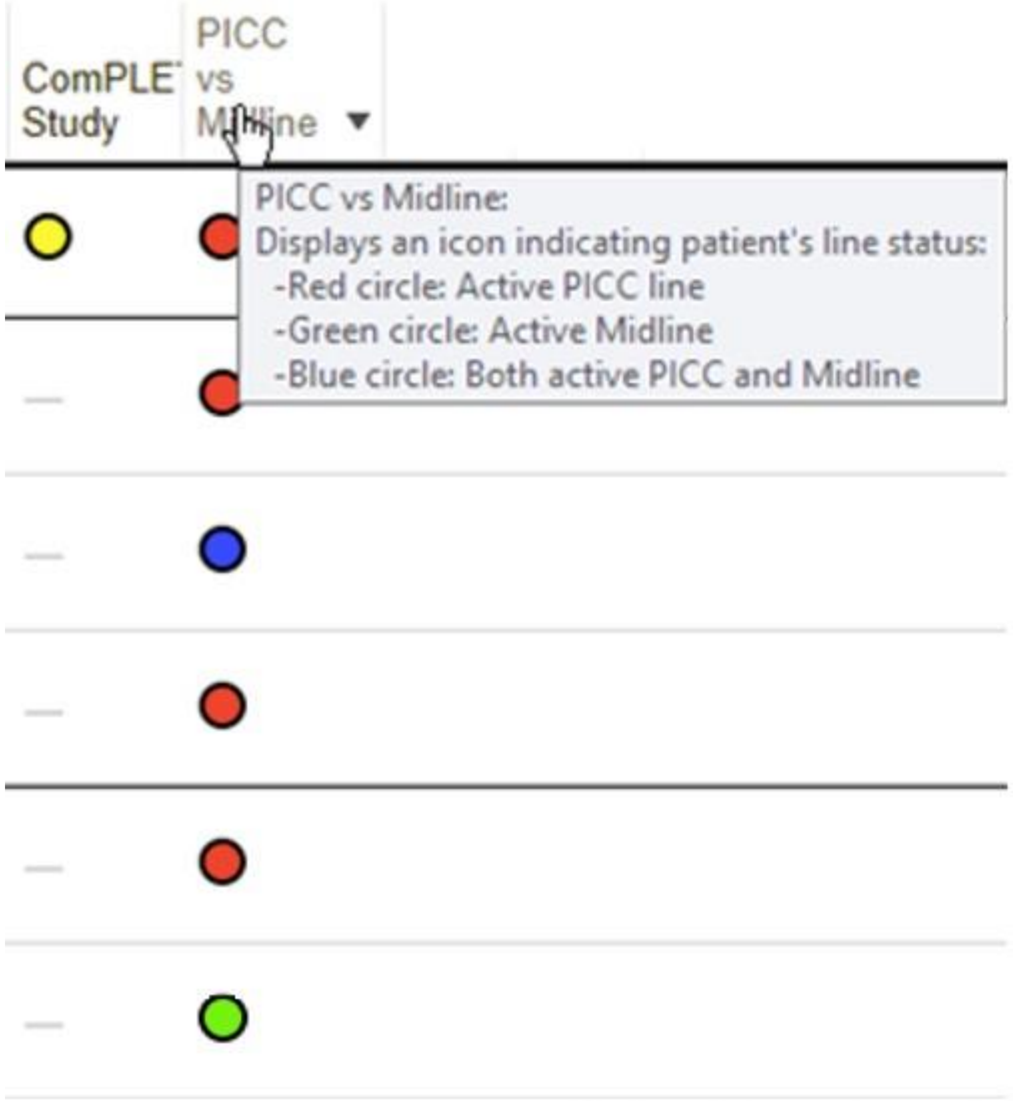


© 2024 Epic Systems Corporation
