## Supplementary figures and images for "Comparing Midline and Peripherally Inserted Central Catheters – a Randomized Feasibility Trial"

### Supplemental Figure 3

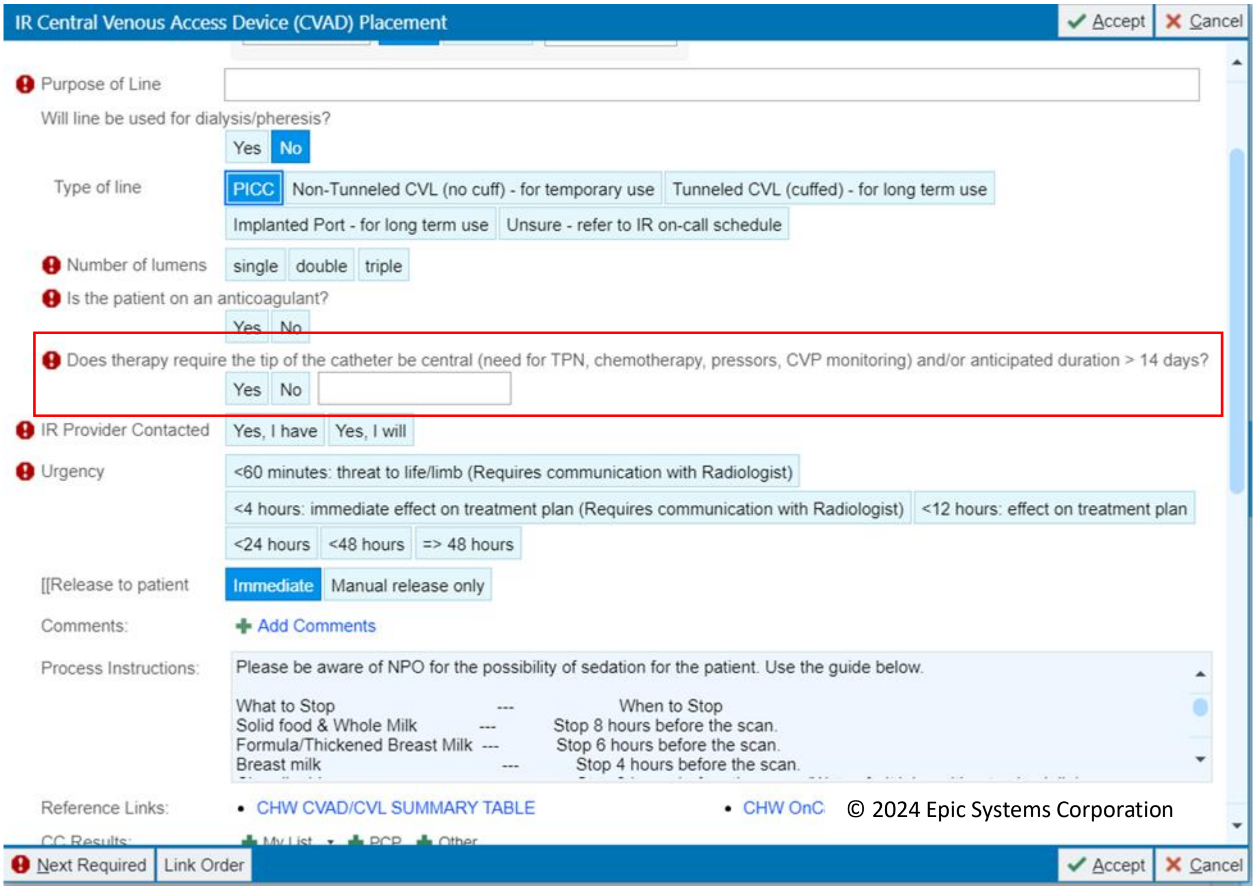


Supplemental Figure 3. PICC order with screening question highlighted in red box.
